# Proteomic Signatures of Conversion Risk and Disease Severity in Multiple Sclerosis

**DOI:** 10.64898/2026.03.25.26348613

**Authors:** Laure Bastide, Virginie Imbault, Gaetano Perrotta, Serena Borrelli, Sophie Elands, Vincent van Pesch, Eva Borràs, Eduard Sabido, Nicolas Gaspard, David Communi, Xavier Bisteau

**Affiliations:** Department of Neurologie, Hôpital Universitaire de Bruxelles, Université Libre de Bruxelles, Brussels, Belgium; Department of Neurology, CHU Nîmes, Université Montpellier, Nîmes, France; IRIBHM J.E. Dumont, Université Libre de Bruxelles ULB, Campus Erasme, Brussels, Belgium; Neuroinflammation Imaging Lab (NIL), Institute of NeuroScience, Université catholique de Louvain, Brussels, Belgium; Laboratory of Neurochemistry, Institute of Neuroscience, Université Catholique de Louvain (UCLouvain), 1200 Brussels, Belgium; Department of Neurology, Cliniques Universitaires Saint-Luc, Université catholique de Louvain (UCLouvain), 1200 Brussels, Belgium; Department of Neurology, Hôpitaux Iris Sud, Brussels, Belgium; Proteomics Unit, Centre de Regulació Genòmica (CRG), Dr. Aiguader 88, 08003 Barcelona, Spain; Proteomics Unit, Universitat Pompeu Fabra (UPF), Dr. Aiguader 88, 08003 Barcelona, Spain; Neurology Department, Yale School of Medecine, New Haven, CT USA

**Author notes:** <u>Correspondance to:</u> Laure Bastide, Erasme Hôpital Universitaire de Bruxelles, Université Libre de Bruxelles, Route de Lennik 808, 1070 Brussels, Belgium; and Xavier Bisteau, IRIBHM J.E. Dumont, Université Libre de Bruxelles ULB, Campus Erasme, Route de Lennik 808, Brussels, Belgium.

**Keywords:** Proteomics, multiple sclerosis, biomarkers signature, cerebrospinal fluid, SWATH-MS

## Abstract

Despite important advances in understanding the etiopathology of multiple sclerosis, factors determining disease progression remain partially understood and often difficult to predict. Specific diagnostic and prognostic biomarkers are needed to optimize the risk-benefit ratio of treatment for each patient. The aim of our study was to identify a cerebrospinal fluid proteomic signature associated with diagnosis and short- to mid-term prognosis across the multiple sclerosis continuum.

Our multicentric cohort study analyzed CSF samples from 120 patients using a proteomics data-independent acquisition strategy. Differentially expressed proteins were identified across diagnostic groups: 62 patients with multiple sclerosis, 15 patients with clinically isolated syndrome, and 43 healthy controls. We also compared the CSF of patients with no evidence of disease activity with those with disease activity at 2 and 5 years of follow-up. A diagnostic and prognostic classification model was built using iterative cross-validated logistic regression models on shared differentially expressed proteins across these two comparisons.

A total of 1,257 proteins were quantified, and 162 differentially expressed proteins were identified across comparisons. We identified a set of ten proteins associated with the diagnosis and prognosis of multiple sclerosis, including previously identified potential biomarkers (CH3L2, IGHG1, IGKC, LAMP2, ADA2), proteins known to be involved in the pathophysiology of multiple sclerosis (A0A8J8YUT9, AT2A2, CO3A1) and two yet unreported proteins (DSC2 and MMRN2). Multivariate models based on these proteins achieved good accuracy for the diagnosis of MS compared with CIS (area under the receiver operating characteristics curve [AUROC] up to 80% using 3 proteins) and prognosis (NEDA vs. EDA; AUROC up to 96% at 2 and 5 years; using 5 proteins).

These results, which will require further investigation to validate the new biomarkers, open new perspectives on multiple sclerosis pathophysiology and therapeutic targets.

## Introduction

Multiple sclerosis is a complex and heterogeneous disease driven by two parallel pathogenic events, a progressive degenerative process superimposed by acute activities of inflammation leading to clinical and MRI relapses and accumulated disability.^1^

Recently, the classification of multiple sclerosis based on three discrete categories characterized by different clinical courses—relapsing-remitting (RRMS), primary progressive (PPMS), and secondary progressive (SPMS)—has evolved toward a spectrum of manifestations, starting with prodromal radiologically isolated syndrome (RIS), a first clinically isolated syndrome (CIS) and evolution to relapsing and progressive multiple sclerosis. This spectrum is thought to be underpinned by the variable expression since the beginning of these two parallel mechanisms, neurodegeneration and inflammation, which are also modulated by patient-specific factors, such as sex, age, social and environmental exposures, genetics, and disease duration.^2^ The newly published 2024 revisions of the McDonald criteria allow the diagnosis of multiple sclerosis in patients presenting with RIS and have unified the framework for the diagnosis of relapsing and progressive forms.^3^ Despite this important progress in understanding multiple sclerosis etiopathology, the course of the disease remains partially understood and poorly predictable. A RIS cohort study showed a cumulative probability of a first clinical event is only 51.2% at 10 years of follow-up.^4^ Furthermore, according to several CIS cohort studies—prior to the revision of the McDonald criteria in 2017—17% to 25% of patients diagnosed with CIS with a normal brain MRI will develop multiple sclerosis within 5 to 20 years of follow-up and with highly variable severity.^5–9^ At the same time, many disease-modifying therapies (DMT) have emerged over the past two decades and different lines of treatment are available depending on the aggressiveness of the disease.^10,11^ Their effectiveness has been reported to be greater on reducing the course of the disease if they are initiated early during the disease progression as soon as CIS or even RIS state, delaying the appearance of disability by years.^12–14^ Specific biomarkers are still needed to better diagnose but also refine the prognosis and predict the disease progression, helping clinicians to optimize the risk-benefit of treatment to each patient.

Several studies have sought to identify biomarkers in blood or CSF for a clearer and earlier diagnosis of multiple sclerosis as well as to more accurately establish the prognosis. Multiple proteins have emerged as biomarkers, such as kappa free light chain (KFLC),^15–17^ neurofilament light chain^18^ or chitinase-3-like 1 protein (CH3L1).^19^ Some of these are currently recommended for use in clinical routine but none are multiple sclerosis-specific.

Clinical proteomics has emerged as an important approach for biomarker discovery in MS, aiding in diagnosis, prognosis, elucidating etiopathogenesis, and monitoring treatment response. Although several technologies are used for proteomic analyses, liquid chromatography coupled with tandem mass spectrometry has established itself over the years as the method of choice for the identification and quantification of proteins.^20,21^ Progress over the last decade in instrument sensitivity, speed of acquisition, and data analysis has enabled the accurate and sensitive identification and quantification of proteins with increasingly deep proteome coverage across large-scale sample sets. Among mass spectrometry strategies, data-independent acquisition (DIA) enables systematic fragmentation of all precursor ions across the full mass range, thereby reducing the stochasticity inherent to data-dependent acquisition (DDA) methods and improving quantitative reproducibility.^22^ Sequential windowed acquisition of all theoretical fragment ion spectra (SWATH-MS) is one of the most widely implemented DIA approaches and has become an essential unbiased and consistently reproducible strategy predominantly used in clinical proteomics focused on neurological diseases; various protocols or tutorials have been developed.^23–35^ This unbiased but highly specific protein identification proteomics approach makes SWATH-MS the ideal tool for diseases as complex as multiple sclerosis, when searching for a unique biomarker, as it allows the identification and quantification of a protein signature.

In this study, we identified proteomic signatures for the diagnosis of multiple sclerosis and the prognosis stratification at the onset, short-term and mid-term using SWATH-MS data acquisition.

## Materials and methods

### Study design and population

The first cohort comprised 69 retrospectively identified patients (31 with multiple sclerosis, 10 with CIS, and 28 controls [CTL]) with leftover CSF samples collected between July 2014 and June 2017 and archived in the Hôpital Universitaire de Bruxelles (HUB) Neurologie/Neurochirurgie biobank (Reference B2014/001). The second cohort comprised 51 prospectively identified patients (31 with multiple sclerosis, 5 with CIS and 15 CTL) with additional CSF collection for research purposes. Samples were collected between March 2017 and December 2022 from patients enrolled in the study when evaluated for suspected multiple sclerosis in one of the three participating departments of neurology in Brussels, Belgium: Departments of Neurology of HUB, Centre Hospitalier Universitaire Brugmann (CHUB), and Cliniques Universitaires de Saint-Luc (CUSL).

The inclusion criteria were as follows: (i) age > 18 years, (ii) availability of lumbar puncture and MRI data, and (iii) ability to provide informed consent.

Patients fulfilling the 2017 McDonald criteria^36^ were designated as having MS or CIS if they did not fulfil the McDonald 2017 criteria for dissemination in space or in time and had no conversion to MS at the end of follow-up; they were excluded if they had another neurological disease diagnosis. All individuals designated as having CTL had a diagnosis of a non-inflammatory neurological disease, CSF oligoclonal bands (OCBs) were absent, and CSF white blood cell count (WCC) was < 3/mm^3^. The clinical characteristics of the CTL group are presented in Supplementary Table S1.

### Clinical follow-up and variables

At lumbar puncture (T0) patients underwent a standardized diagnostic evaluation including the Expanded Disability Status Scale (EDSS), brain and spinal MRIs, followed by regular clinical and MRI follow-up.

Time to clinical or MRI relapse (TTR) was defined as the time elapsed from treatment initiation to the occurrence of the first clinically confirmed relapse or MRI relapse, defined as a new T2 lesion on spinal or brain MRI.

Annualized clinical or MRI relapse rate (ARR) calculated as the ratio of the previously described TTR to the follow-up in years.

No Evidence of Disease Activity (NEDA) was defined as follows: (1) no clinical relapse, (2) no disability worsening (based on EDSS progression) and (3) no MRI activity (new T2 lesion or Gd+ lesion or enlarging T2 lesion), according to the NEDA-3 criteria^37^ at 2 years and 5 years after the initiation of treatment in MS patients or the relapse onset for CIS patients.

The age-related MS severity score (ARMSS) was calculated by adjusting the EDSS scores for the patient’s age at the time of assessment. Factoring age in severity assessment allows cross-sectional analysis of patient-level disability data.^38^

Descriptive statistics were performed on the demographic and clinical variables; two-sided Fisher’s exact test or chi-squared test were used for contingency tables or two-sided Mann–Whitney U-test and Kruskal–Wallis’ test were used for continuous variables, as appropriate. A correction for multiple comparisons was performed using the Benjamini–Hochberg method (*FDR*-value).

### Ethics statement

This study was reviewed and approved by the Ethics Committee of Erasme Hopital Universitaire de Bruxelles under reference B406201733112-P2017/417. The participants provided written informed consent to participate in the prospective study.

### Sample collection and handling

CSF samples were collected during the initial diagnostic assessment before intravenous steroid administration if needed, when possible, or any DMT. Retrospective leftover CSF samples were stored at -20°C whereas prospective CSF samples were processed according to published guidelines.^39^

CSFs were thawed and concentrated using a centrifugation column (Centricon 3kDa, Millipore Merck) at 4°C and 14.000g until 25 to 30µL.

The depletion of the top 14 most abundant proteins, corresponding to albumin and immunoglobulins, was performed using affinity columns (Pierce Top 14 Abundant Protein Depletion Spin Columns; Thermo Scientific), as described by the manufacturer. Protein concentration in the samples was measured after depletion using the Folin method.^40^ Proteins were diluted in 25mM ammonium carbonate and dithiothreitol was added to obtain a final concentration of 25mM. Each sample was incubated for 1 h at 4°C. Alkylation of samples was performed by the addition of iodoacetamide with final concentration of 71mM before incubation for 1h at 4°C. All samples were diluted 4 times with acetone and incubated à - 20°C during 1h for protein precipitation. After centrifugation at 13.000 rpm for 20 min at 4°C, acetone was removed and 40 ngr of trypsin (Gold Mass Spectrometry Grade V5280 Promega, USA WI) was added. Samples were incubated and shaked at 37°C during 30 min and then over night without shaking. Samples were incubated and shaked at 37°C during 30min and then over night without shaking.

Finally, the pH of each sample was adjusted to 2 with a 10% formic acid (FA) solution. Samples were then cleaned up using a 1 mL high-lipophilic balance column equilibrated with acetonitrile (ACN) 5%/0.1% FA, eluted with ACN 80%/0.1% FA, dried in a vacuum concentrator, and resuspended in buffer A (H2O 100%/HCOOH 0.1%).

### Data dependent analysis

Proteins were identified from CSF samples using label-free DDA mass spectrometry (TripleTOF 5600 SWATH, AB Sciex) coupled with micro-high-performance liquid chromatography (µLC 425, Eksigent). Peptide separation was performed by applying a 90 min gradient hydrophobic separation on a column (ChromXP C18 CL, 150 mm x 0.3 mm, 3 µM, 120 A, Sciex) using a two-step ACN gradient with FA: 5–25% ACN / 0.1% FA in 48 min then 25%–60% ACN / 0.1% FA in 20 min and were sprayed online in the mass spectrometer. Subsequently, peptides were ionized by electrospray ionization and injected into the TripleTOF 5600 mass spectrometer (TT 5600, AB Sciex). The 20 most intense precursors with charge states of 2 to 4 were selected for fragmentation. MS1 spectra were collected in the range of 400–1250 m/z with an accumulation time of 250 ms, and MS2 spectra were collected in the range of 100–2000 m/z for 100 ms.

### Generation of spectral libraries

The generated .WIFF files were directly analysed by ProteinPilot software (AB Sciex) and the recorded spectra were matched to peptides by the search algorithm Paragon (AB Sciex) using the database human Swiss-Prot part of UniProtKB.^41^ Trypsin was selected as digestion enzyme. Oxidation at methionine was set as a dynamic modification and carbamidomethylation as a static modification at cysteine. A decoy database search was performed with a target false discovery rate of < 1%.

### Data independent analysis

1 µg of peptides was injected using SWATH-MS acquisition on a Triple TOF 5600 mass spectrometer (Sciex, Concord, Canada) interfaced to an Eksigent NanoLC Ultra 2D HPLC System (Eksignet, Dublin, CA). Peptides were injected on a separation column (Eksigent ChromXP C18 CL, 150 mm x 0.3 mm, 3 µM, 120 A, Sciex) using a two steps acetonitrile gradient (5%–25% ACN / 0.1% HCOOH in 48 min then 25%–60% ACN / 0.1% HCOOH in 20 min) and were sprayed online into the mass spectrometer. Swath acquisitions were performed using 42 windows of a fixed effective isolation width to cover a mass range of 200–1500 m/z. SWATH MS2 spectra were collected from 200–1500 m/z. The collision energy for each window was determined according to the calculation for a charge 2+ ion centered upon the window with a spread of 15. An accumulation time of 76 ms was used for all fragment-ion scans in high-sensitivity mode and for the survey scans in high-resolution mode acquired at the beginning of each cycle, resulting in a duty cycle of ∼3.3 s.

Raw data were converted to mzML format with MSConvert (Version 3.0) and their processing was carried out with DIA-NN^42^ (version 1.8.2 beta 27) with search parameters set as follows: precursor *FDR* 1%; mass accuracy at MS1 and MS2 both set to 0; scan window set to 0; isotopologues and MBR turned on; protein inference at gene level; heuristic protein inference enabled; quantification strategy set to QuantUMS (high accuracy); neural network classifier double-pass mode; cross-run normalization RT-dependent. A universal library was used, and protein re-annotation was performed. Spectra were searched against a prior *in loco* generated library containing 9972 proteins covered by 174348550 peptides including for common contaminants.^43^

To generate the library, DDA and DIA data acquired on TT5600 were used to generate one universal library by FragPipe^44^ (version 18.0) using a pre-defined workflow DIA_SpecLib_Quant. Specifically, decoys were first added to the FASTA which contains human protein sequences (UP000005640, 78986 reviewed and unreviewed entries, downloaded from UniProtKB on 07^th^ June 2022). Then, MSFragger (version 3.5) was used to search DDA raw data, with the following settings: precursor and fragment mass tolerance 30□ppm; strict trypsin with no more than five missed cleavages; peptide length 6–50; peptide mass 500–5000; C□+□57.021464 as fixed modification; M□+□15.9949, N-term +42.0106, nQ -17.0265, nE -18.0106, DN +0.984016 and carbamylation +43.005814 as variable modifications; min matched fragments 4; max fragment charge 2. In the validation step, MSBooster was implemented on both spectra and RT levels, and then Percolator and ProteinProphet integrated in Philosopher (version 4.4.0) were used for PSM validation and protein inference. Library generation was conducted using EasyPQP (version 0.1.50) with RT calibration based on ciRTs and Lowess fraction set to 0.04. Only fragment types b and y were included with a tolerance of 15□ppm with a max delta_unimod.

### Differential expression analysis

MSstats (version 4.16.1)^45^ in R (version 4.5.1)^46^ was used to perform the data processing, model-based estimation of the quantity of each protein based on a relative log_2_ transformation, and group comparison following the workflow described previously.^47^ All samples underwent median normalization. Only the top 100 features were selected for protein summarization, uninformative features and outliers were removed. No specific peptides were excluded, and proteins with only one feature or with features present in less than three measurements per run were excluded. No missing value imputation was performed. For the group comparison analysis, we selected proteins present in at least 90% of the runs in at least one group of comparisons. The group comparison was performed between MS vs CTL, MSCIS vs CTL, CIS vs CTL, MS vs CIS, EDA vs NEDA at 2 and 5 years of follow-up. A correction for multiple comparisons was performed using the Benjamini–Hochberg method, and a threshold for adjusted *P-value* < 0.05 referred to as false discovery rate (*FDR*) in the figures or table was deemed significant.

### Diagnostic and prognostic classification models

A four-fold cross-validation was performed to identify the most discriminative combination of proteins, as previously described.^48^ Patient data were divided into four equal subgroups in the training set. For each group, each protein was fitted on a logistic regression model between the two groups, and its classification ability was evaluated using the area under the receiver operating characteristics (AUROC) curve. The most discriminative protein was selected as the first classifier. Additional discriminative proteins were repeatedly added while increasing AUROC values. The proteins selected at least twice within the four cross-validation steps in the training set were chosen as the characteristic classification signature for that training set. The best classification signature for each training set was fitted on a logistic regression model and applied to the validation set. The procedure from dividing the data into training and validation sets to fitting the logistic model with the best classification signature was repeated 500 times to assess the reproducibility of the classification ability. A final consensus model was comprised of the combination of proteins that were selected most frequently in the 500 repeats. To obtain the upper level for the predictive accuracy of the selected consensus proteins, the final model was fitted to the full dataset, and the predictive accuracy was quantified using the AUROC, sensitivity, specificity, and accuracy. The estimate of variability associated with the receiver operating characteristics (ROC) curve was obtained by plotting the 25th and the 75th quantile of the sensitivities for each value of 1-specificity obtained in the validation set over all the iterations for which each protein combination was selected. The pROC package in R was used to draw ROCs, calculate AUROCs, and other performance measures (*i.e.,* sensitivity, specificity, and accuracy).

For each classification task, we identified the top three most frequently selected protein combinations, representing the most robust predictors across resampling. ROC curves of the top three selected combinations in each classification case are graphed (solid lines), representing performance on the full cohort and reported with their AUROC and confidence interval (CI) at 95% among the entire cohort. Dotted lines show the distribution of the ROC curves across the 500 validation iterations. To evaluate whether CSF proteomic signatures could predict short-and mid-term inflammatory activity (NEDA vs. EDA at 2 and 5 years) in multiple sclerosis, we applied the same iterative, cross-validated logistic regression framework used for diagnostic analyses.

To assess whether simpler protein signatures could retain strong discriminative power, we evaluated the performance of multivariate logistic models constructed using the unique proteins contributing to the top three combinations of each comparison. These models were evaluated on the entire cohort using bootstrap resampling and are depicted by the purple solid line.

## Results

### Differential clinical profiles of CIS and MS groups inform biomarker identification strategy

A total of 187 samples were initially considered. As outlined in Supplementary data Fig. S1, three samples failed the initial quality control, and 184 samples were processed using mass spectrometry. The second quality filter based on protein identification depth and signal intensity led to the exclusion of an additional 64 samples. Ultimately, 120 samples fulfilled all analytical requirements and were retained for biostatistical analyses, comprising 69 retrospective samples and 51 prospective samples (17 from HUB, 14 from CHUB, and 20 from CUSL). Of those 62 had MS, 15 had CIS and 43 were CTL.

The clinical characteristics of MS, CIS, and CTL patients at last follow-up are presented in Table 1 and statistics are reported in Supplementary File S1.

**Table 1.** Baseline and clinical characteristics of the patients with multiple sclerosis, clinically isolated syndrome and controls.

| Clinical characteristics of the cohort |  |  | MS | CIS | CTL | FDR-value* | MS multi-ple scler-osis pa-tients, CIS clinically iso-lated syn-drom e pa-tients, CTL con-trols pa-tients, CSF cere-bro-spinal fluid, WCC white cells count, OCBs Oligo-go-clonal bands, ON optic neur-i-tis, TM trans-verse mye-litis, PF pos-terior fos-sae, Enc en-cepha-litic MRI |
| --- | --- | --- | --- | --- | --- | --- | --- |
| **Cohort size** |  |  | **62** | **15** | **43** |  |  |
| Number of MS and CIS diagnosis at T0 | *n* |  | 59 | 18 |  |  |  |
| Number of conversion CIS to MS | *n* |  |  | 3 |  |  |  |
| Delay onset to diagnosis | Days Median (range) |  | 60.5 (2–748) | 30 (7–1095) |  | 0.463 |  |
| Age<sup>a</sup> | Years Median (range) |  | 30 (14–80) | 35 (19–55) | 38 (20–82) | **0.045** |  |
| Sex | F / M |  | 46 / 16 | 12 / 3 | 24 / 19 | 0.129 |  |
| Follow-up | Years Median (range) |  | 4 (0–10) | 3.14 (2–10) | NA | 0.705 |  |
| CSF data<sup>a</sup> | WCC | Median (range) | 4 (0–59) | 3 (0–17) | 1 (0–4.2) | **< 0.001** |  |
|  | OCBs | Yes / No | 59 / 3 | 9 / 6 | 0 / 40<sup>b</sup> | **< 0.001** |  |
| Clinical onset | ON | *n* (%) | 9 (15%) | 11 (73%) |  | **< 0.001** |  |
|  | TM | *n* (%) | 28 (46%) | 1 (7%) |  | **0.014** |  |
|  | PF | *n* (%) | 14 (23%) | 1 (7%) |  | 0.352 |  |
|  | Enc | *n* (%) | 10 (16%) | 2 (13%) |  | 1 |  |
| Annualized clinical and MRI relapse rate |  |  | 0.18 (0–2.35) | 0 |  | **< 0.001** |  |
| Number of clinical relapse | Median (range) |  | 0 (0–2) | 0 |  | **0.020** |  |
| Time to MRI or clinical relapse after treatment start | Months |  | 12.1 (-1.1–93.5) | NA |  | NA |  |
| EDSS | Baseline | Median (range) | 2 (0–7.5) | 2 (0–3.5) |  | 0.585 |  |
|  | End | Median (range) | 1.5 (0–8) | 1 (-3) |  | 0.176 |  |
| MRI data | Brain T2 lesion | 0 lesion | 0 | 8 (53%) |  | **< 0.001** |  |
|  |  | 1 to 3 lesions | 9 (15%) | 5 (33%) |  | 0.250 |  |
|  |  | 4 to 9 lesions | 23 (37%) | 2 (13%) |  | 0.176 |  |
|  |  | > 10 lesions or confluent | 30 (48%) | 0 |  | **< 0.001** |  |
|  | Spinal T2 lesion | No | 15 (26%) | 12 (80%) |  | **< 0.001** |  |
|  |  | Yes | 46 (77%) | 3 (20%) |  | **< 0.001** |  |
|  | Gd+ lesions | No | 25 (40%) | 11 (73%) |  | 0.067 |  |
|  |  | Yes | 37 (60%) | 4 (27%) |  | 0.067 |  |
| Treatment baseline / End<sup>b</sup> |  | First line | 42 / 30 | 0 / 1 |  | **< 0.001 / < 0.001** |  |
|  |  | Second line | 5 / 18 | 0 / 0 |  | 0.661 / **0.030** |  |
|  |  | None | 11 / 10 | 15 / 14 |  | **< 0.001 / < 0.001** |  |
| NEDA at 2y *n* = 60<sup>b</sup> | Yes / no |  | 21 / 26 | 13 / 0 |  | **< 0.001** |  |
| NEDA at 5y *n* = 44<sup>b</sup> | Yes / no |  | 4 / 34 | 6 / 0 |  | **0.038** |  |
| ARMSS at the end of FU<sup>b</sup> | Median (range) |  | 3 (0.3–8.9) | 1 (0.4–5) |  | **0.030** |  |

magnetic resonance imagery, EDSS expanded disability status scale, Gd+ gadolinium enhancement, NEDA no evidence of disease activity by no relapses, no increase in disability and no new or active lesion on MRI.
\*P was calculated for MS vs CIS except for age, sex and CSF data it was MS vs CIS vs CTL patients. Two-sided Fisher's exact test or Chi squared test when appropriate were used for contingency tables or two-sided Mann–Whitney U-test and Kruskal–Wallis's test when appropriate for continuous variables. A correction for multiple comparisons was done by Benjamini–Hochberg method (FDR-value).
<sup>a</sup>Age was significantly different only between MS and CTL patients. CSF white cells count was significantly different between MS vs CTL and CIS vs CTL patients but not between MS vs CIS. OCBs were significantly different between MS vs CIS patients.
<sup>b</sup> Missing data exists. Three CTL patients had no OCBs information. Treatment baseline and end (*n* = 58); For MS patients were lost of follow-up before starting a treatment during and one did not benefit DMT because of an ongoing cancer treatment and was classify as none DMT. ARMSS (*n* = 61).

MS patients were significantly older than CTL individuals (30 [14–80] vs. 38 [20–82], *FDR*-value = 0.025). CSF OCBs were more frequent in MS than in CIS patients (95% vs. 60%, *FDR*-value < 0.001). Clinical onset differed substantially: optic neuritis being the presenting syndrome in 73% of CIS patients versus 15% of MS patients (*FDR*- value < 0.001).

Over half of the CIS patients (53%) had no brain lesions at baseline, whereas all MS patients presented with at least one lesion. None of the CIS patients exhibited more than 10 brain lesions, in contrast to 48% of the MS patients. Seventy seven percent of the MS patients presented with at least one spinal lesion on MRI, whereas this was observed in only 20% of the CIS patients. Although not statistically significant, MS patients tended to show a higher proportion of gadolinium-enhancing lesions (60% vs 27%, *FDR*-value = 0.067). Almost no CIS patients had received DMT at baseline. Markers of inflammatory disease activity, including the number of clinical relapses and annualized relapse rate (ARR) were all higher in MS than in CIS (0 [0–2] vs. 0, *FDR*-value = 0.020 and 0.18 [0–2.35] vs 0, *FDR*-value < 0.001 respectively). Measures of disability progression (EDSS) showed limited discriminatory power, but the ARMSS score was significantly higher in MS patients (3 [0.3–8.9] vs. 1 [0.4–5], *FDR*-value = 0.030). Forty five percent of MS patients achieved NEDA at 2 years and 11% at 5 years.

### CSF proteomic differences reflect disease stage and activity across the CIS–multiple sclerosis continuum

We performed an in-depth CSF proteomic analysis combining DDA- and DIA/SWATH-MS on 120 clinically well-characterized samples. We quantified 1,257 proteins, of which 593 were selected for downstream analysis, as described in the Methods section.

Considering multiple sclerosis as a continuum that begins prior to the CIS stage, we structured our analyses to compare the combined MS and CIS group (MSCIS) against CTL, complemented by separate comparisons of MS vs CTL and CIS vs CTL to identify potential diagnostic biomarkers. Building on the clinical differences distinguishing CIS from MS, as well as the NEDA outcomes, we also searched for differentially expressed proteins associated with prognosis at disease onset and at short- and mid-term follow-up. For this purpose, differential expression analysis was performed between MS and CIS patients, and within the overall MSCIS cohort according to NEDA status at 2 and 5 years as illustrated in Figure 1 and reported in Suppelmentary File S2.

**Figure 1.**
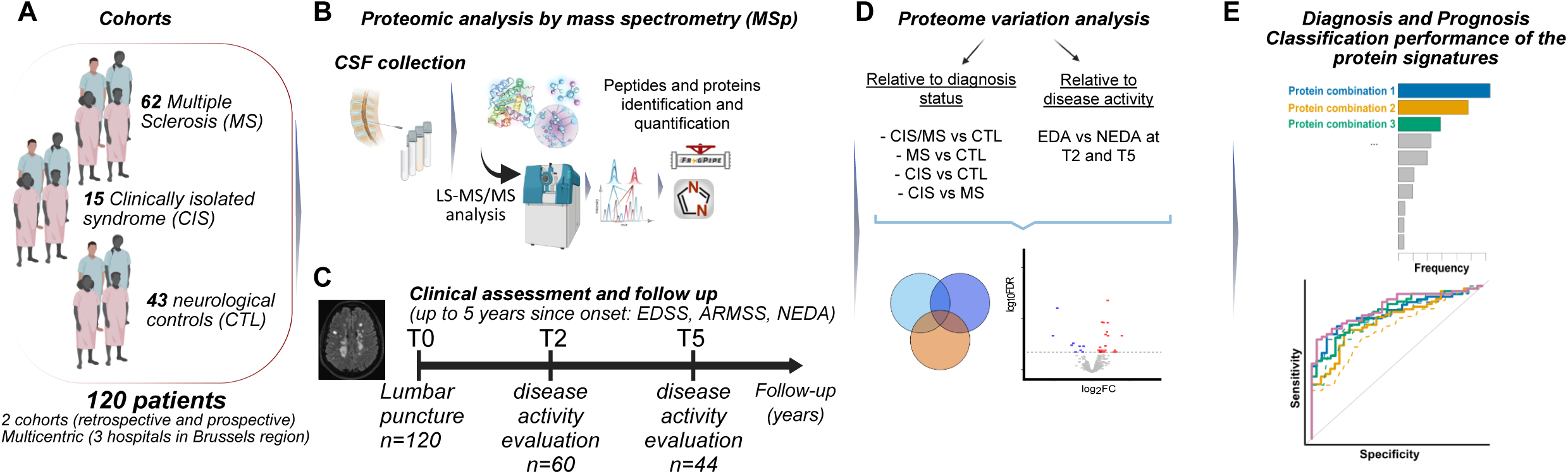
Experimental design of the study. **(A)** Retrospective and prospective enrollment of multiple sclerosis (MS), clinically isolated syndrome (CIS) and controls (CTL) patients form 3 Belgium sites Hopital Universitaire de Bruxelles (HUB), Centre Hospitalier Universitaire de Brugmann (CHUB) and Cliniques Universitaires de Saint-Luc (CUSL). **(B)** Preparation and acquisition of the collected CSF samples by label-free data independent acquisition. **(C)** Clinical follow-up of MS and CIS patients up to five years. **(D)** Differential expression analysis performed with the MSstats R-package across diagnostic groups (MS vs. CTL, MSCIS vs. CTL, CIS vs. CTL and MS vs. CIS) and disease activity vs. no evidence of disease activity at 2 and 5 years of follow-up in the MSCIS group. **(E)** Diagnosis and prognosis performance of the 15 shared proteins of the 6 groups’ comparisons evaluated by logistic regression model with cross-validation.

Differential expression analysis revealed distinct proteomic signatures between MS, CIS, CTL, NEDA vs. EDA at 2 and 5 years. In total, 53 proteins were differentially expressed between MS and CTL, 32 between MSCIS and CTL, 13 between CIS and CTL and 4 between MS and CIS (Fig. 2A). The largest foldchanges were observed between MS and CTL. No differentially expressed protein was common to the three comparisons of a single categories (MS vs CTL, CIS vs CTL and MS vs CIS), only one overlapped between the MS vs CIS and CIS vs CTL comparisons, two between MS vs CTL and MS vs CIS and seven (12%) between MS vs CTL and CIS vs CTL (Fig. 2B). In contrast, as many as 18 differentially expressed proteins overlapped between comparison of disease activity (NEDA status) at 2 and 5 years (Fig. 2B).

**Figure 2.**
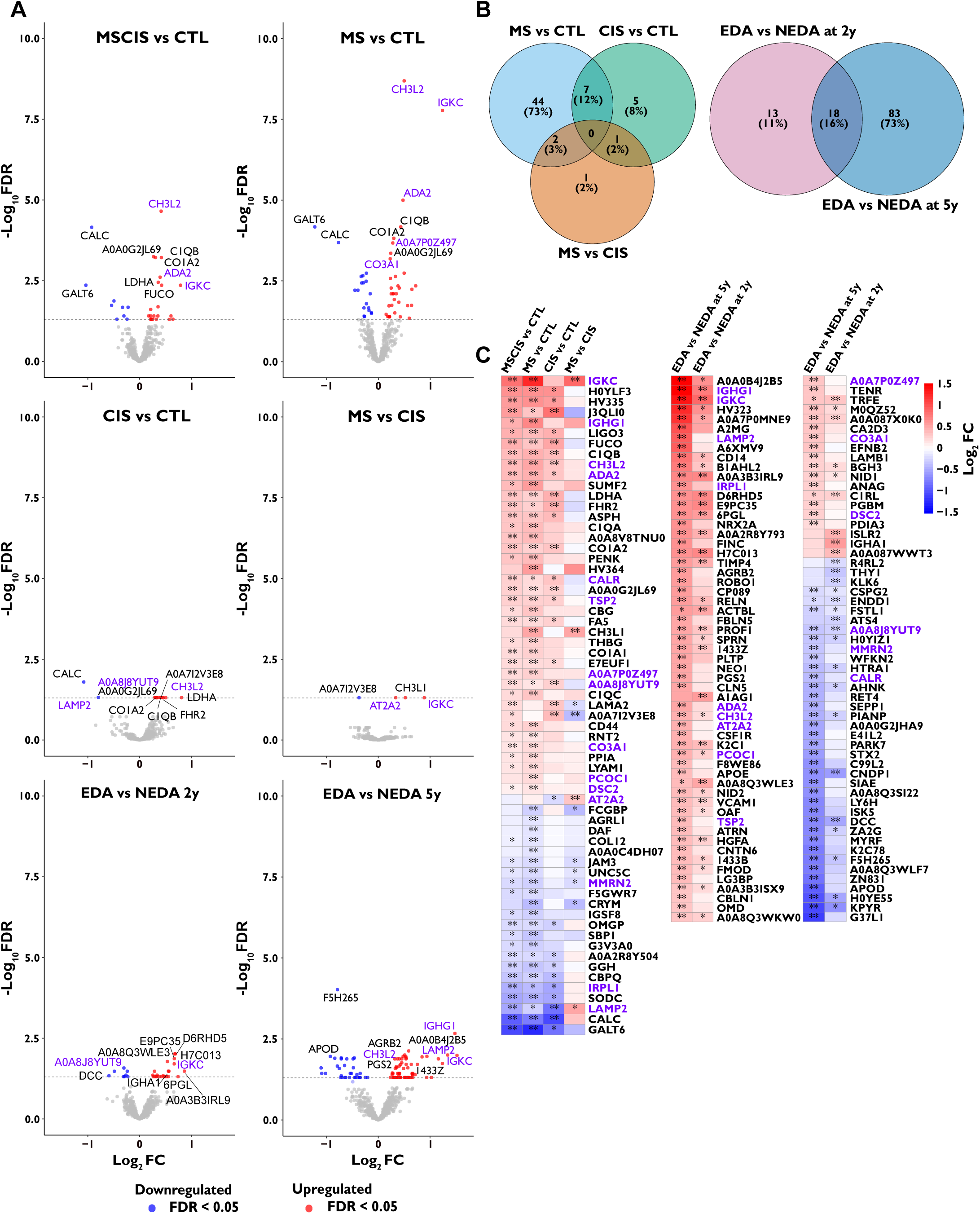
Overview of the proteome variations in relation to compared groups of patients. Differential expression analysis of CSF proteins in MS and CIS (*n* = 77) patients versus CTL subjects (*n* = 43) and among MS (*n* = 62) and CIS (*n* = 15) patients between active (*n* = 26/34) versus stable disease (*n* = 34/10) defined by the NEDA achievement at 2 and 5 years of follow-up. **(A)** Volcano plots showing differentially expressed proteins with a false discovery rate (*FDR*) < 0.05 in relation to the compared groups. The differential expression analysis was performed by the MSstats R-package based on linear mixed-effects models. The top 10 upregulated or downregulated proteins are marked. The proteins in purple are common differentially expressed proteins across the 6 comparisons. **(B)** Venn diagrams highlighting the proteins overlap between the different groups’ comparison. **(C)** Heatmaps based on the log_2_ fold change (FC) of all identified differentially expressed proteins grouped by condition comparisons and activity vs stable disease at 2 and 5 years of follow-up and showing their behavior in other groups comparisons. The asterisk represents the level of statistical significance: ** *FDR* < 0.05, * *P* < 0.05.

To visualize the relationships of these protein signatures across all comparisons, heatmaps in the Figure 2C displayed how the proteins identified in each comparison behave in others comparisons, revealing clusters of proteins that are tightly associated with specific disease states or progression profiles (Fig. 2C).

Altogether, the proteomic analysis of CSF from MS and CIS patients compared with CTL individuals revealed diagnostic specificity, minimal overlap at early disease stages, and greater prognostic consistency across the CIS–multiple sclerosis continuum.

### Identification of integrated biomarkers correlating with disease progression and initial spinal cord pathology

To better understand which molecular changes are shared between early multiple sclerosis stages and mid-term disease activity, we examined the overlap between proteins identified in the diagnostic comparisons (MS, CIS, CTL) and those associated with disease activity (NEDA vs EDA at 2 and 5 years). This revealed 15 proteins common to both diagnostic and prognostic contrasts (Fig. 3A). These proteins represent a core set of candidates that are not only dysregulated early in the CIS-multiple sclerosis continuum but also remain associated with short and mid-term disease activity, making them suitable candidates for further investigation.

**Figure 3.**
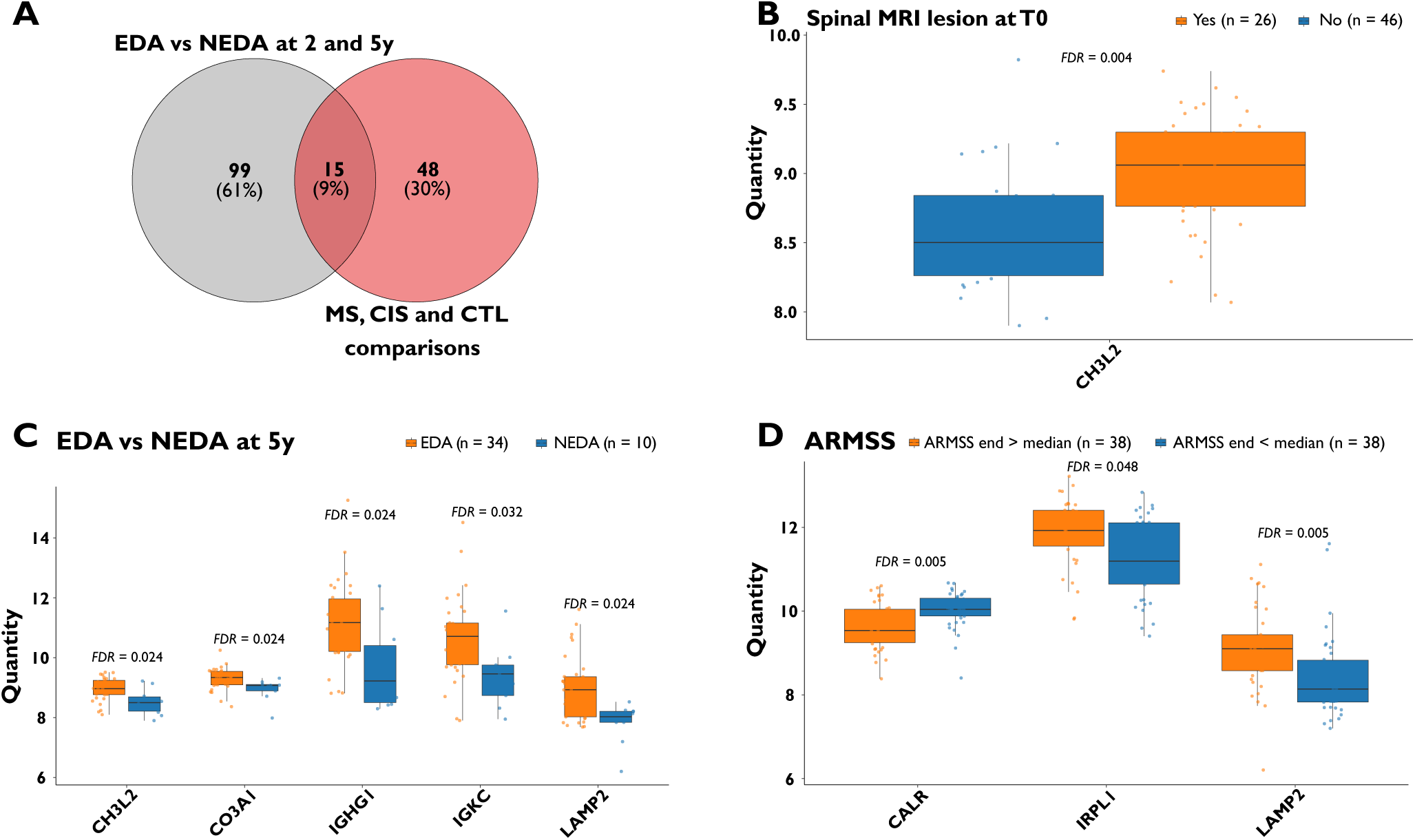
Identification of common differentially expressed proteins between diagnostic and prognostic groups. **(A)** Venn diagram showing the overlap between both groups’ comparison: NEDA vs EDA representing all the identified proteins of the active vs stable disease at 2 and 5 years of follow-up and MS, CIS and CTL comparisons grouping the differentially expressed proteins of the 4 conditions’ comparisons. Boxplots of the statistically relevant association assessed by a two-sided Mann–Whitney U-test corrected for multiple comparison with Benjamini–Hochberg method between these 15 proteins and **(B)** the presence or absence of spinal MRI lesion at the initial assessment, **(C)** ARMSS at the end of follow-up compared to the median (2.45 [0.3-8.9]) of the MSCIS group, and **(D)** the EDA versus NEDA at 5 years of follow-up. The boxplot represents the median, Q1, Q3 and the IQR.

Among these 15 shared proteins summarized in the Table 2, several have already been described in the literature as potential diagnostic or prognostic biomarkers in multiple sclerosis. These include A0A7P0Z497,^49^ ADA2,^50,51^ CH3L2,^52^ CALR^53,54^ and LAMP2.^55^ Other proteins in this group have recognized roles in multiple sclerosis pathophysiology: for example, A0A8J8YUT9 belongs to a receptor family involved in immune-mediated neuroprotective interactions,^56^ and CO3A1 is strongly induced in both active and inactive multiple sclerosis brain lesions.^57^ AT2A2, another protein in this set, is known to be modified by dimethyl fumarate (DMF), a commonly used DMT in multiple sclerosis.^58^ We also confirmed dysregulation of classic humoral immune markers such as IGHG1 and IGKC consistent with intrathecal B-cell activation and KFLC which is a fragment of IGKC, already used in the clinical routine and observed in multiple sclerosis. Importantly, we identified five proteins (DSC2, IRPL1, MMRN2, PCOC1, TSP2) for which no evidence has yet been reported in the multiple sclerosis literature, highlighting novel candidates that may contribute to early disease mechanisms or progression.

**Table 2.** Description of the common 15 differentially expressed proteins between conditions MS-CIS-CTL and active versus stable disease comparisons.

| Common proteins | Protein name | Previously described as potential bio-marker | Reference |
| --- | --- | --- | --- |
| **A0A7P0Z497** | Peptidyl-prolyl cis-trans isomerase | No, but elevated in MS patients and in EAE mice | Fan et al. 2024 |
| **A0A8J8YUT9** | Tyrosine-protein kinase receptor | No, but the receptor family is involved in immune-mediated neuroprotective interactions in MS | Stadelmann et al. 2002 |
| **ADA2** | Adenosine deaminase 2 | Yes, potential biomarker of the active phase of MS | Hinsinger et al. 2024 |
| **AT2A2** | Sarcoplasmic/endoplasmic reticulum calcium ATPase 2 | No, but modified by the dimethyl fumarate | Hermann et al. 2019 |
| **CALR** | Calreticulin | Elevated in SPMS compared to control<br>Increased in sera of MS patients compared to control | Hejazi et al. 2024<br>Ní Fhlathartaigh et al. 2013 |
| **CH3L2** | Chitinase-3-like protein 2 | Yes, for diagnosis and prognosis | Mollgaard et al. 2016 |
| **CO3A1** | Collagen alpha-1(III) chain | No, but its gene is strongly induced in active and even more in inactive MS lesions | Mohan et al. 2010 |
| **DSC2** | Desmocollin-2 | No |  |
| **IGHG1** | Immunoglobulin heavy constant gamma 1 | Yes, locally synthesized IgG in the CSF of MS patients consists mainly of IgG1<br>Serum IgG1 can discriminate between MS vs control, SPMS vs RRMS and SPMS vs PPMS | Kashka et al. 1979<br>Zhou et al. 2023 |
| **IGKC** | Immunoglobulin kappa constant | Yes, its fragment KFLC for diagnosis of MS and prognosis. | Levrault et al. 2022<br>Levrault et al. 2023 |
| **IRPL1** | Interleukin-1 receptor accessory protein-like 1 | No |  |
| **LAMP2** | Lysosome-associated membrane glycoprotein 2 | Yes, predictor of the MS progression in serum | Xu et al. 2023 |
| **MMRN2** | Multimerin-2 | No |  |
| **PCOC1** | Procollagen C-endopeptidase enhancer 1 | No |  |
| **TSP2** | Thrombospondin-2 | No |  |

Table summarizing the 15 proteins with their complete name and if they were already described as potential biomarkers in scientific literature.

We next evaluated whether the abundance of these proteins correlated with baseline clinical features, including age, sex, clinical onset, MRI T2 lesion burden in the brain and spinal cord, gadolinium enhancement and OCB presence. CH3L2 was significantly increased in MSCIS patients presenting at least one spinal lesion at baseline (9.061 [8.763–9.299] vs. 8.501 [8.262–8.841], *FDR* = 0.004; Fig. 3B), suggesting a link with early spinal cord involvement.

We then assessed the association of these proteins with markers of disease severity and progression. Proteins were examined against the EDSS and ARMSS scores at the end of the follow-up, and the disease inflammatory activity with TTR, ARR, and NEDA status at 2 and 5 years. CALR levels were decreased in patients with a higher-than-median ARMSS score (9.539 [9.247–10.044] vs. 10.044 [9.886–10.309], *FDR* = 0.005; Fig. 3C), while IRPL1 and LAMP2 were elevated in the same subgroup (11.928 [11.553–12.409] vs. 11.195 [10.646– 12.108], *FDR* = 0.048 and 9.1 [8.578–9.437] vs. 8.138 [7.828–8.825], *FDR* = 0.005, respectively), suggesting potential links with disability progression.

In addition, CH3L2, CO3A1, IGHG1, IGKC and LAMP2 were all higher in patients who experienced disease activity at 5 years, compared with those who maintained NEDA (respectively 8.966 [8.768–9.245] vs. 8.501 [8.217–8.693], *FDR*-value = 0.024, 9.340 [9.096–9.542] vs. 9.076 [8.892–9.105], *FDR*-value = 0.024, 11.172 [10.208–11.961] vs. 9.221 [8.501–10.406], *FDR*-value = 0.024, 10.714 [9.768–11.156] vs. 9.457 [8.740–9.753], *FDR*-value = 0.032, 8.928 [8.027–9.366] vs. 8.028 [7.844–8.210], *FDR*-value = 0.024; Fig. 3D). These findings underscore the prognostic relevance of this protein subset and its potential role in predicting long-term disease activity.

### Iterative cross-validated regression reveals highly discriminative CSF proteomic signatures along the CIS–multiple sclerosis continuum

To evaluate the diagnostic potential of the 15 proteins shared between diagnostic and prognostic comparisons, we applied the logistic regression framework combined with a fourfold cross-validation, repeated over 500 iterations (see the Methods section for further description).

For the MSCIS vs CTL classification (Fig. 4A), CH3L2 emerged as the most recurrently selected protein (113 selections), achieving an AUROC of 0.762 (*P* = 8×10□□) (Fig. 4A). The combination of CH3L2 and MMRN2 was selected 89 times and performed better (AUROC 0.809, *P* = 4×10□□). ADA2, selected 75 times, also showed strong discriminative ability (AUROC 0.748, *P* = 1×10□□). The results were similar when focusing on MS vs CTL (Fig. 4B). The most frequently selected combination, CH3L2 and MMRN2 (126 selections), achieved an AUROC of 0.832 (*P* = 2×10□□). CH3L2 alone (96 selections) reached an AUROC of 0.783 (*P* = 5×10□□), while the combination CH3L2 + IGKC (58 selections) yielded an AUROC of 0.831 (*P* = 9×10□□). These results highlight CH3L2 as a central diagnostic protein across early multiple sclerosis phenotypes.

**Figure 4.**
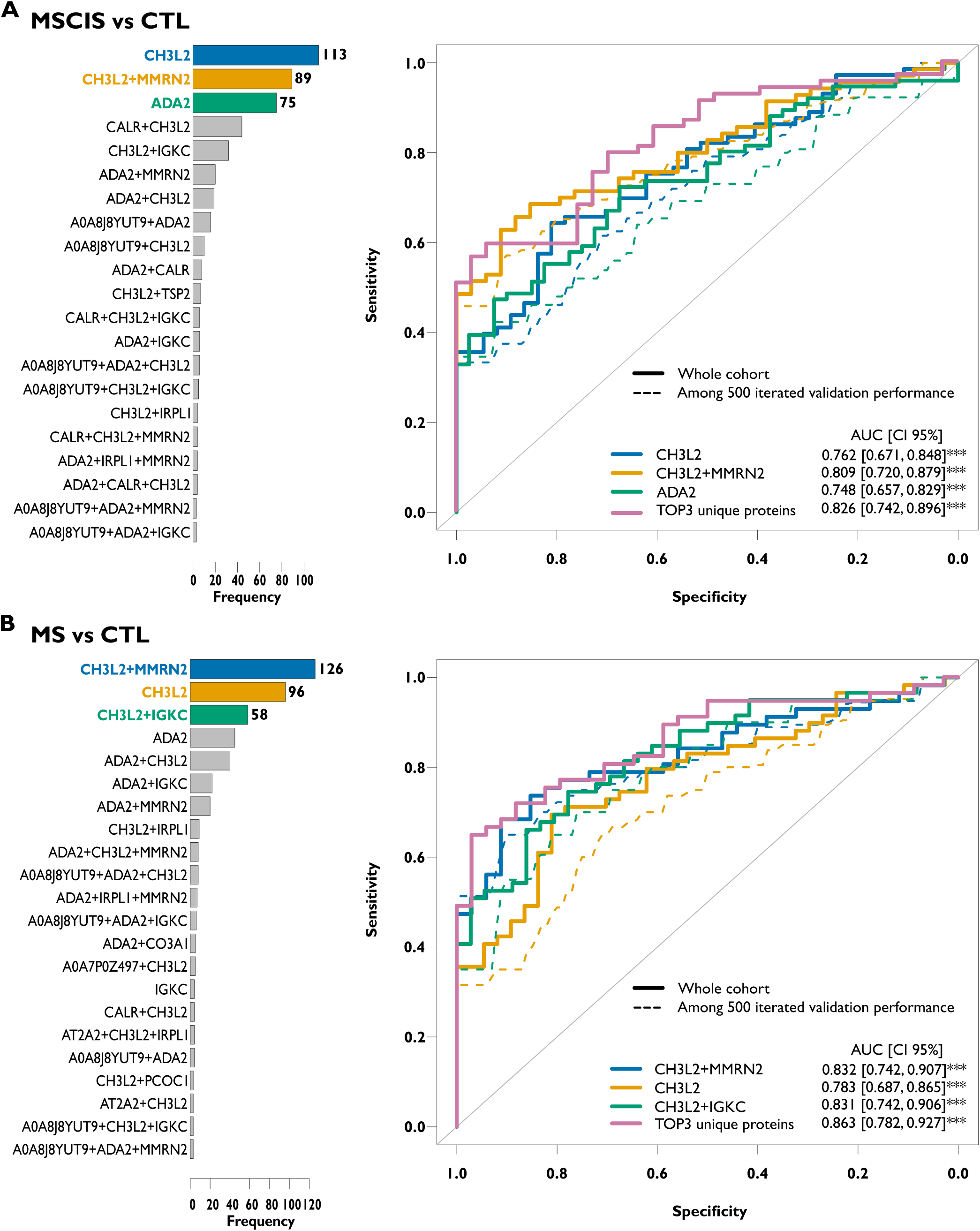
MSCIS vs CTL and MS vs CTL classification performance of the 15 common proteins. Performance of the 15 proteins for predicting diagnosis of MSCIS vs CTL and MS vs CTL determined by a logistic regression model which uses the area under the receiver operating characteristics curve (AUROC) to assess the predictive performance of a protein, with a forward selection of the proteins and a cross validation by splitting the cohort in training/validation groups (2/3-1/3) over 500 iterations. **(A)** Frequency plots represent the top three protein combination that was the most selected as the best classifier over the 500 iterations. **(B)** Predictive power of the top three protein combination in differentiating MSCIS (*n* = 77) vs CTL (*n* = 43) and MS (*n* = 62) vs CTL (*n* = 43), is analyzed by AUROC. The AUROC scores were assessed by a two-sided Mann–Whitney U-test. The *P* were in the order: 8×10^-6^, 4×10^-7^, 1×10^-5^, 2×10^-7^, 5×10^-6^ and 9×10^-8^. The purple AUROC was obtained with a multivariate logistic regression with bootstrapping on the entire cohort for the unique proteins from the top three protein combinations (TOP3 unique proteins). The *P* were in the order 1×10^-7^ and 1×10^-8^

For the CIS vs CTL comparison (Fig. 5A), the combination CH3L2 + LAMP2 (44 selections) provided an AUROC of 0.824 (*P* = 0.0003). The combination ADA2 + AT2A2 (41 selections) slightly outperformed it (AUROC 0.837, *P* = 0.0002), while A0A8J8YUT9 + ADA2 (30 selections) also performed well (AUROC 0.816, *P* = 0.0004). These combinations suggest that CIS, although clinically milder, already harbors detectable proteomic deviations from CTL.

**Figure 5.**
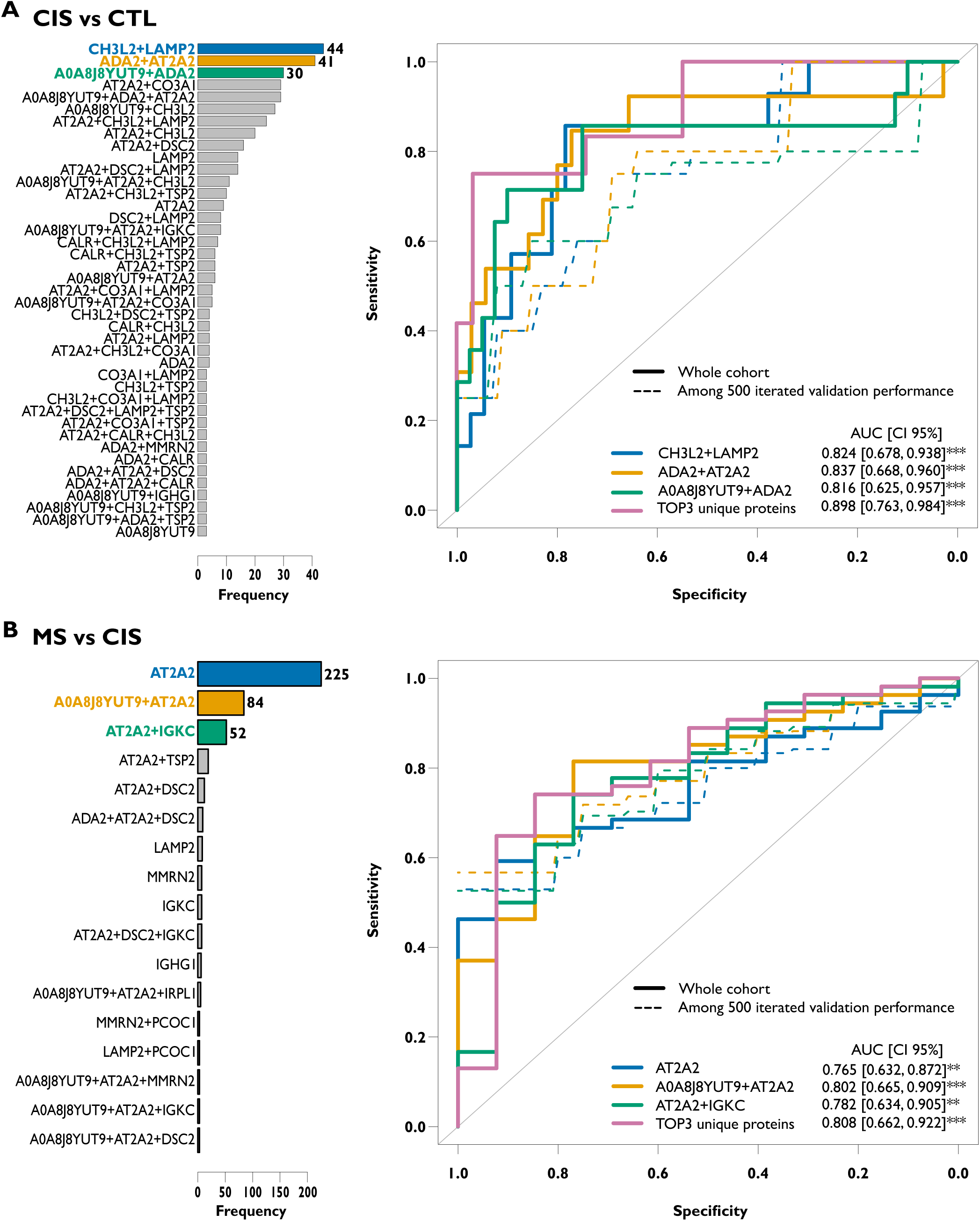
CIS vs CTL and MS vs CIS classification performance of the 15 common proteins. Performance of the 15 proteins for predicting diagnosis of CIS vs CTL and MS vs CIS determined by a logistic regression model which uses the area under the receiver operating characteristics curve (AUROC) to assess the predictive performance of a protein, with a forward selection of the proteins and a cross validation by splitting the cohort in training/validation groups (2/3-1/3) over 500 iterations. **(A)** Frequency plots represent the top three protein combination that was the most selected as the best classifier over the 500 iterations. **(B)** Predictive power of the top three protein combination in differentiating CIS (*n* = 15) vs CTL (*n* = 43) and MS (*n* = 62) vs CIS (*n* = 15), is analyzed by AUROC. The AUROC scores were assessed by a two-sided Mann–Whitney U-test. The *P* were in the order: 0.0003, 0.0002, 0.0004, 0.004, 0.0009 and 0.002. The purple AUROC was obtained with a multivariate logistic regression with bootstrapping on the entire cohort for the unique proteins from the top three protein combinations (TOP3 unique proteins). The *P* were in the order 2×10^-5^ and 0.0007.

Finally, the MS vs CIS classification (Fig. 5B) revealed AT2A2 as the most robust discriminator (225 selections, AUROC 0.765, *P* = 0.004). The combinations AT2A2 + A0A8J8YUT9 (84 selections, AUROC 0.802, *P* = 0.0009) and AT2A2 + IGKC (52 selections, AUROC 0.782, *P* = 0.002) further improved classification, underscoring AT2A2 as a key protein distinguishing early MS from CIS.

To assess whether simpler protein signatures could retain strong discriminative power, we next evaluated the performance of multivariate logistic models constructed using the unique proteins contributing to the top three combinations of each comparison. These models were evaluated on the entire cohort using bootstrap resampling (Figures 4 and 5).

The multi-protein diagnostic signatures demonstrated strong and consistent performance across all comparisons. For distinguishing MSCIS from CTL (Fig. 4A), the three-protein combination CH3L2, MMRN2 and ADA2 achieved an AUROC of 0.826 (*P* = 1×10□□). In the MS vs CTL classification (Fig. 4B), the signature composed of CH3L2, MMRN2 and IGKC performed even better, reaching an AUROC of 0.863 (*P* = 1×10□□). The highest diagnostic accuracy was observed for CIS vs CTL, where a five-protein panel (CH3L2, LAMP2, ADA2, AT2A2 and A0A8J8YUT9) yielded an AUROC of 0.898 (*P* = 2×10□□).

Finally, the ability to discriminate MS from CIS (Fig. 5A) was supported by the combination AT2A2, A0A8J8YUT9 and IGKC, which reached an AUROC of 0.808 (*P* = 0.0007). Collectively, these results demonstrate that multiple protein combinations, particularly those involving CH3L2, MMRN2, ADA2, IGKC, AT2A2 and LAMP2, provide strong and reproducible diagnostic power across the MS-CIS-CTL comparisons. These findings highlight a set of highly informative proteins capable of differentiating early multiple sclerosis phenotypes and controls with high accuracy.

### Proteomic profiling of CSF reveals signatures that predict short-and mid-term inflammatory activity in multiple sclerosis

Focusing on comparisons between patients with EDA and those achieving NEDA, we identified a restricted set of proteins that consistently contributed to prognostic discrimination at both 2 and 5 years of follow-up (Fig. 6).

**Figure 6.**
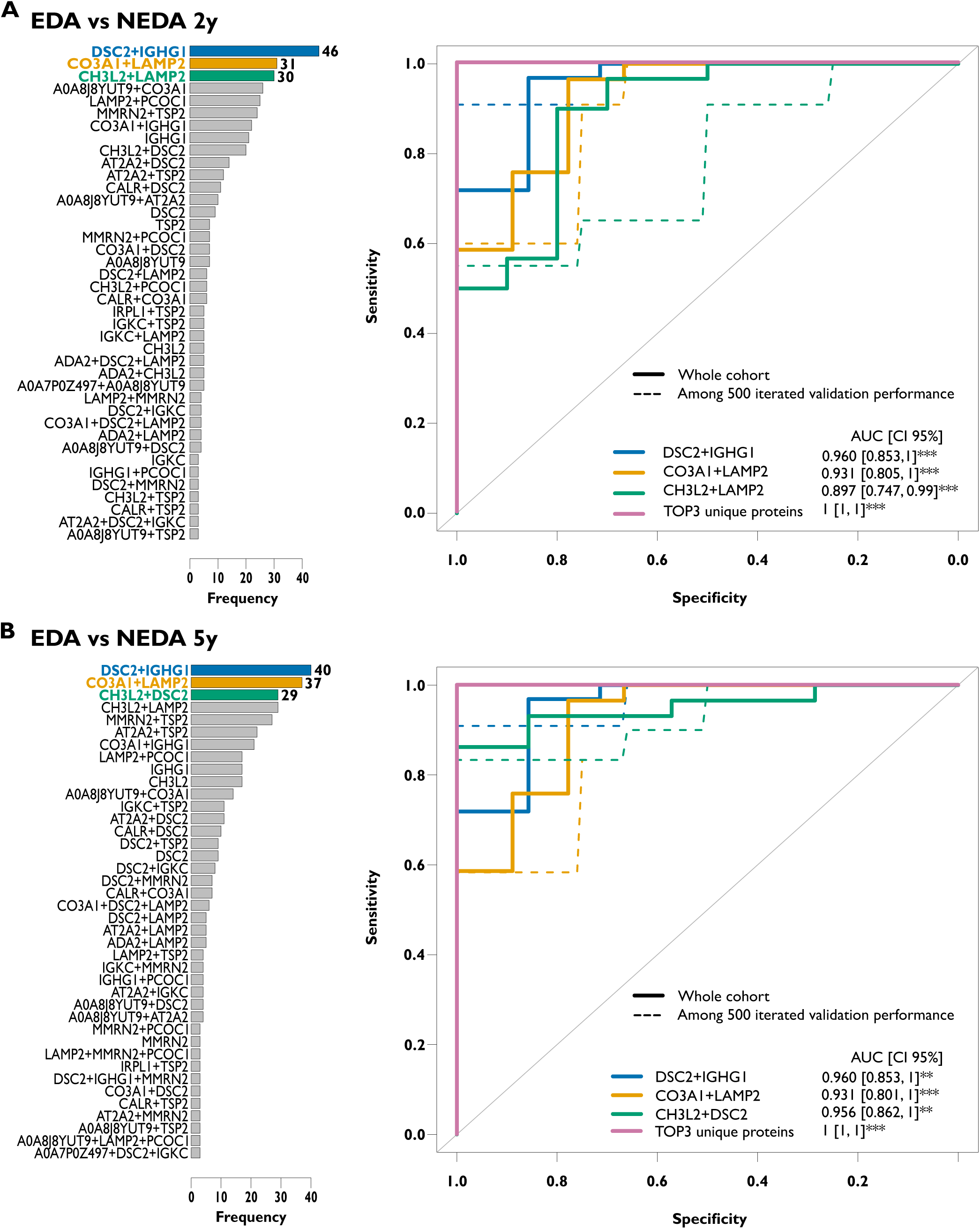
NEDA vs EDA classification performance of the 15 common proteins at 2 and 5 years of follow-up. Performance of the 15 proteins for predicting prognosis by the disease activity at 2 and 5 years of follow-up after treatment start determined by a logistic regression model which use the area under the receiver operating characteristics curve (AUROC) to assess the predictive performance of a protein, with a forward selection of the proteins and a cross validation by splitting the cohort in training/validation groups (2/3-1/3) over 500 iterations. **(A)** Frequency plots represent the top three protein combination that was the most selected as the best classifier over the 500 iterations. **(B)** Predictive power of the top three protein combination in differentiating EDA (*n* =26) vs NEDA (*n* = 34) at 2 years and EDA (*n* = 34) vs NEDA (*n* = 10) at 5 years, is analyzed by AUROC. The AUROC scores were assessed by a two-sided Mann-Whitney U-test. The *P* were in the order: 8×10^-4^, 3×10^-4^, 4×10^-5^, 2×10^-5^, 3×10^-5^, and 3×10^-5^. The purple AUROC was obtained with a multivariate logistic regression with bootstrapping on the entire cohort for the unique proteins from the top three protein combinations (TOP3 unique proteins).

At 2 years (Fig. 6A), the three most frequently selected protein combinations across 500 cross-validation iterations were DSC2 + IGHG1 (46 occurrences), which achieved the highest performance with an AUROC of 0.960 (*P* = 2×10□□), followed by CO3A1 + LAMP2 (31 occurrences, AUROC 0.931, *P* = 3×10□□), and CH3L2 + LAMP2 (30 occurrences, AUROC 0.897, *P* = 8×10□□). These results reveal a strong and coherent prognostic signal, with multiple combinations converging on proteins associated with immune activity, extracellular matrix remodeling and inflammatory signaling.

At 5 years, the two leading combinations were identical to those identified at 2 years; DSC2 + IGHG1 (40 occurrences, AUROC 0.960, *P* = 2×10□□) and CO3A1 + LAMP2 (37 occurrences, AUROC 0.931, *P* = 3×10□□), demonstrating remarkable longitudinal stability of the prognostic proteomic substrate. The third-ranked combination differed only slightly, with CH3L2 + DSC2 emerging as a robust discriminator (29 occurrences, AUROC 0.956, *P* = 3×10□□).

To further evaluate the prognostic value of these markers, we aggregated the unique proteins from the top combinations at 2 and 5 years and generated a multivariate logistic regression model. The signature combining DSC2, IGHG1, CO3A1, LAMP2 and CH3L2, yielded an AUROC of 1.0 for discriminating between EDA and NEDA in the full dataset at 2 and 5 years.

## Discussion

In this study, we took advantage of the strengths of the SWATH-MS approach—its reproducible quantification and broad proteomic coverage—to analyze 120 CSF samples and identify predictive proteomic signatures associated with diagnosis, onset and both short- and mid-term prognosis in early multiple sclerosis. Across all comparisons, we quantified 1,257 proteins and identified 162 differentially expressed proteins.

The comparison between MS and CTL showed the higher number and amplitude of changes in diagnostic comparisons, consistent with a more pronounced inflammatory and neurodegenerative substrate in established multiple sclerosis. Despite the biological overlap between CIS and multiple sclerosis (heatmap Fig. 2C), very few DEPs were shared across diagnostic categories. This limited overlap underscores the heterogeneity and stage-specific molecular alterations across early demyelinating presentations and highlights the potential for proteins to reflect distinct biological processes in CIS versus MS groups.

We observed a higher overlap when comparing EDA vs NEDA at 2 and 5 years. This suggests that biomarkers associated with inflammatory activity and clinical or MRI progression may be more stable and consistently detectable over time than those discriminating diagnostic categories.

Distinct expression patterns emerge for multiple sclerosis-specific markers, CIS-driven proteins, and those linked to disease activity, supporting the idea that different biological axes (diagnosis vs prognosis) rely on partially non-overlapping molecular pathways.

Among the identified differentially expressed proteins, 15 were common to diagnostic and prognostic group comparisons. Within these proteins, CH3L2 was significantly elevated in patients with at least baseline spinal T2 lesions, whereas LAMP2 was higher in patients with greater disability progression. When assessing the diagnostic and prognostic potential of these 15 shared proteins, 10 proteins were identified with strong discriminative ability across comparisons. ADA2, AT2A2, A0A8J8YUT9, CH3L2, CO3A1, IGHG1, IGKC and LAMP2 contributed most consistently to diagnostic classification, whereas DSC2, IGHG1, CO3A1, LAMP2 and CH3L2—alone or in combination—demonstrated excellent discrimination between EDA and NEDA at both time points. As expected, such performance reflects an overfitting inherent to the aggregation of all top-ranked proteins and highlights the need for external validation. Nonetheless, the reproducibility of DSC2, IGHG1, CO3A1 and LAMP2 across complementary analyses and time points strongly supports their relevance as candidate prognostic biomarkers. This recurrence could also underscore their shared involvement in the biological processes associated with disease activity.

Chitinase-3-like protein 2 (CH3L2), is a well-documented biomarker for MS. Expressed by astrocytes and microglia, CH3L2 has been associated with the conversion of CIS to multiple sclerosis, shorter relapse-free intervals and disability progression.^52,59,60^ In our analysis, CH3L2 appeared in both diagnostic signatures (MSCIS vs CTL, MS vs CTL, CIS vs CTL) and prognostic models differentiating EDA from NEDA-3 at T2 and T5, in agreement with existing literature, although we did not detect significant differences between MS and CIS.

Immunoglobulins are central to multiple sclerosis pathophysiology and are established biomarkers, notably through OCBs^36^ and KFLC.^3,15–17,61,62^ As KFLC derives from IGKC, its presence in our diagnostic and prognostic signatures aligns with previous findings. IGHG1, the constant region of IgG1, has recently been implicated in multiple sclerosis pathogenesis: plasma IgG1 aggregates induce complement-dependent apoptosis in neurons and astrocytes and discriminate multiple sclerosis from healthy control (HC) or other neurological disease controls with high accuracy (AUC 0.93).^63^ IgG1 also differentiates SPMS from RRMS and PPMS (AUC 0.91–0.92).^64^ IGHG1 and IGKC genes were also highly expressed in all lesion types in the brain white matter of patients with progressive multiple sclerosis.^65^ These data strongly support the biological plausibility of our IGHG1-based prognostic models.

Lysosome-associated membrane glycoprotein 2 (LAMP2) plays a critical role in autophagy, a pathway implicated in multiple sclerosis pathogenesis.^66^ A previous study reported elevated serum LAMP2 in SPMS vs RRMS, though CSF LAMP2 lacked diagnostic utility for distinguishing multiple sclerosis from HC.^55^ In our cohort, CSF LAMP2 levels were higher in patients with ARMSS progression and in those experiencing EDA at 5 years; LAMP2 also contributed to the top prognostic signatures for EDA vs NEDA-3 at both T2 and T5. Another study investigating iron metabolism disorders in multiple sclerosis identified LAMP2 within a multi-gene predictive model, showing that its expression in both blood and brain samples helped distinguish MS patients from controls.^67^ In our cohort, CSF LAMP2 did not contribute to the classification of MS or MSCIS versus CTL, although it appeared among the top three protein combinations distinguishing CIS from CTL. Notably, a third publication examining X-linked genetic factors associated with multiple sclerosis susceptibility reported 20 SNP-linked genes that were significantly more abundant in MS patients, and LAMP2 was among them.^68^ Consistent with this, LAMP2 up-regulation in MS group was also described in pediatric multiple sclerosis.^69^

Adenosine deaminase 2 (ADA2) regulates extracellular adenosine, a molecule involved in neurotransmission, inflammation, regeneration, and blood brain barrier (BBB) permeability.^70^ Of interest, the purine nucleoside analogue 2-chlorodeoxyadenosine (cladribine), a treatment for active multiple sclerosis, is resistant to deamination by ADA. As a result, its nucleotide derivatives accumulate in lymphocytes and induce CD4□, CD8□ and CD19□ cell death.^71,72^ In line with previous studies, ADA2 was differentially expressed between MS and CTL groups and could discriminate them (AUROC 0.748).^50,51^

For several proteins identified in our signatures, clinical biomarker data are lacking, but mechanistic studies support their involvement in multiple sclerosis. A0A8J8YUT9 is an isoform of NTRK2 which interacts with is involved in CNS homeostasis and multiple sclerosis pathogenesis.^56,73^ CO3A1, encoding collagen alpha-1 (III) chain, is strongly induced in active and inactive multiple sclerosis lesions.^57,65^ AT2A2 regulates cytosolic calcium via SERCA2 activity and modulates T and B cell receptor signaling; its modification by DMF in experimental autoimmune encephalomyelitis mouse models suggests functional relevance.^58^

Finally, our work describes two proteins not previously associated with multiple sclerosis: desmocollin-2 (DSC2) and multimerin-2 (MMRN2). DSC2 is a desmosomal adhesion protein implicated in congenital heart disease^74^ and several cancers.^75,76^ Notably, DSC2 was recently identified as a dominant entry receptor for Epstein-Barr virus (EBV) infection of epithelial cells,^77^ a finding of interest given the strong evidence linking EBV to multiple sclerosis development.^78,79^ MMRN2, an extracellular matrix glycoprotein involved in vascular stability and angiogenesis may be relevant in multiple sclerosis given the prominent role of BBB dysfunction in demyelinating lesions.^80–82^

Our study has limitations, including mixed retrospective and prospective sampling, heterogeneous pre-analytical handling, a limited number of patients with CIS, and the absence of an external validation cohort. To mitigate these issues, we applied volume and peptide normalization prior to mass spectrometry acquisition, selected samples based on protein depth and intensity thresholds, and restricted downstream analyses to proteins detected in at least 90% of runs within a comparison group and we employed cross-validation to strengthen internal robustness. Furthermore, the multicenter nature of the study increased its generalizability.

In summary, our study confirms several proteins as reliable diagnostic and prognostic biomarkers in multiple sclerosis (CH3L2, IGHG1, IGKC, LAMP2, ADA2), identifies additional candidate biomarkers supported by pathophysiological evidence (A0A8J8YUT9, AT2A2, CO3A1), and introduces DSC2 and MMRN2 as novel proteins of interest. Although further validation is required, these findings provide new insights into multiple sclerosis biology and highlight promising avenues for biomarker development and therapeutic exploration.

## Data availability

The mass spectrometry proteomics data have been deposited to the ProteomeXchange Consortium via the PRIDE^83^ partner repository with the dataset identifier PXD073146.

## Acknowledgements

We would like to thank Dr. Serena Borelli and the entire team at the CHUB Neurology Department, as well as Prof. Van Pesch and his team at the Neurology Department of the Cliniques Universitaires de Saint Luc for their collaboration in exchanging CSF samples.

## Supplementary material

Supplementary material is available at *Brain* online.

## Authors contributions

LB and VI handled CSF samples and prepared samples for proteomics analysis. VI performed the MS analysis under the supervision of DC and XB. LB, EB and ES were involved in the data analysis of the SWATH-MS data. LB, GP and NG designed the clinical study. NG, DC and XB were involved in the overall design and supervised the study. LB with the help of GP, SB, SE and VVP recruited patients and compiled all the clinical data used in the study. LB and XB wrote the manuscript and all the authors have read and revised the article and approved the submitted version.

## Supplementary detailed proteomics method

### Samples protocol preparation

CSFs were thawed and concentrated with a centrifugation column (Centricon 3kDa, Millipore Merck) at 4°C and 14.000g until 25 to 30µL.

The depletion of the top 14 most abundant proteins, was performed on affinity columns (Pierce Top 14 Abundant Protein Depletion Spin Columns; Thermo Scientific) then the protein concentration was measured with the Folin method.^1^

Proteins were diluted in 25mM ammonium carbonate and dithiothreitol was added for a final concentration of 25mM. Each sample was incubated during 1h at 4°C. Alkylation of samples was performed by the addition of iodoacetamide with final concentration of 71mM before incubation during 1h at 4°C. All samples were diluted 4 times with acetone and incubated à - 20°C during 1h for protein precipitation. After centrifugation at 13.000 rpm during 20min at 4°C, acetone was removed and 40 ng of trypsin (Gold Mass Spectrometry Grade V5280 Promega, USA WI) was added. Samples were incubated and shaked at 37°C during 30min and then over night without shaking.

The pH of each sample was adjusted at 2 with a 10% formic acid (FA) solution. Finally, the clean-up of the fractions was achieved using a 1mL High Lipophilic Balance column equilibrated by acetonitrile (ACN) 80% and ACN 5%/0.1% FA, eluted by ACN 80%/0.1% FA and dried in a vacuum concentrator and resuspended in tampon A (H20 100%/HCOOH 0.1%).

### Proteomic Analysis of CSF

#### Data Dependant Analysis

Proteins were identified from CSF samples by label-free DDA mass spectrometry (TripleTOF 5600 SWATH, AB Sciex) coupled to micro-high-performance liquid chromatography (µLC 425, Eksigent). Peptides separation was performed by applying a 90min gradient hydrophobic separation on a column (ChromXP C18 CL, 150mm x 0.3mm, 3µM, 120A, Sciex) using a two-step ACN gradient with FA: 5-25% ACN / 0.1% FA in 48min then 25%-60% ACN / 0.1% FA in 20min and were sprayed online in the mass spectrometer. Subsequently, peptides were ionized by electrospray ionization (ESI) and injected into the TripleTOF 5600 mass spectrometer (TT 5600, AB Sciex). The 20 most intense precursors with charge state 2 to 4 were selected for fragmentation. MS1 spectra were collected in the range 400-1250 m/z with an accumulation of 250ms and MS2 spectra were collected in the range 100-2000 m/z for 100ms.

#### Generation of Spectral Libraries

The generated .WIFF files were directly analysed by ProteinPilot software (AB Sciex) and the recorded spectra were matched to peptides by the search algorithm Paragon (AB Sciex) using the database human Swiss-Prot part of UniProtKB.^2^ Trypsin was selected as digestion enzyme. Oxidation at methionine was set as dynamic modification, carbamidomethylation as static modification at cysteine. Decoy database search was performed with target false discovery rate < 1% (FDR).

#### Data Independent Analysis

1µg of peptides was injected using SWATH-MSp acquisition on a Triple TOF 5600 mass spectrometer (Sciex, Concord, Canada) interfaced to an Eksigent NanoLC Ultra 2D HPLC System (Eksignet, Dublin, CA). Peptides were injected on a separation column (Eksigent ChromXP C18 CL, 150mm x 0.3mm, 3µM, 120A, Sciex) using a two steps acetonitrile gradient (5-25% ACN / 0.1% HCOOH in 48min then 25%-60% ACN / 0.1% HCOOH in 20min) and were sprayed online in the mass spectrometer. Swath acquisitions were performed using 42 windows of fix effective isolation width to cover a mass range of 200-1500m/z. The collision energy for each window was determined according to the calculation for a charge 2+ ion centered upon the window with a spread of 15. An accumulation time of 76ms was used for all fragment-ion scans in high-sensitivity mode and for the survey scans in high-resolution mode acquired at the beginning of each cycle, resulting in a duty cycle of ∼3.3s.

Raw data were converted to mzML format with MSConvert (Version 3.0) and their processing was carried out with DIA-NN^3^ (version 1.8.2 beta 27) with search parameters set as follows: precursor FDR 1%; mass accuracy at MS1 and MS2 both set to 0; scan window set to 0; isotopologues and MBR turned on; protein inference at gene level; heuristic protein inference enabled; quantification strategy set to QuantUMS (high accuracy); neural network classifier double-pass mode; cross-run normalization RT-dependent. A universal library was used, and protein re-annotation was performed. Spectra were searched against a prior *in loco* generated library containing 9972 proteins covered by 174348550 peptides including for common contaminants^4^.

To generate the library, DDA and DIA data acquired on TT5600 were used to generate one universal library by FragPipe^5^ (version 18.0) using a pre-defined workflow DIA_SpecLib_Quant. Specifically, decoys were first added to the FASTA which contains human protein sequences (UP000005640, 78986 reviewed and unreviewed entries, downloaded from UniProtKB on 07^th^ June 2022). Then, MSFragger (version 3.5) was used to search DDA raw data, with the following settings: precursor and fragment mass tolerance 30lJppm; strict trypsin with no more than five missed cleavages; peptide length 6–50; peptide mass 500–5000; ClJ+lJ57.021464 as fixed modification; MlJ+lJ15.9949, N-term +42.0106, nQ -17.0265, nE -18.0106, DN +0.984016 and carbamylation +43.005814 as variable modifications; min matched fragments 4; max fragment charge 2. In the validation step, MSBooster was implemented on both spectra and RT levels, and then Percolator and ProteinProphet integrated in Philosopher (version 4.4.0) were used for PSM validation and protein inference. Library generation was conducted using EasyPQP (version 0.1.50) with RT calibration based on ciRTs and Lowess fraction set to 0.04. Only fragment types b and y were included with a tolerance of 15lJppm with a max delta_unimod.

