## Supplementary Figure 1 for "Proteomic Signatures of Conversion Risk and Disease Severity in Multiple Sclerosis"

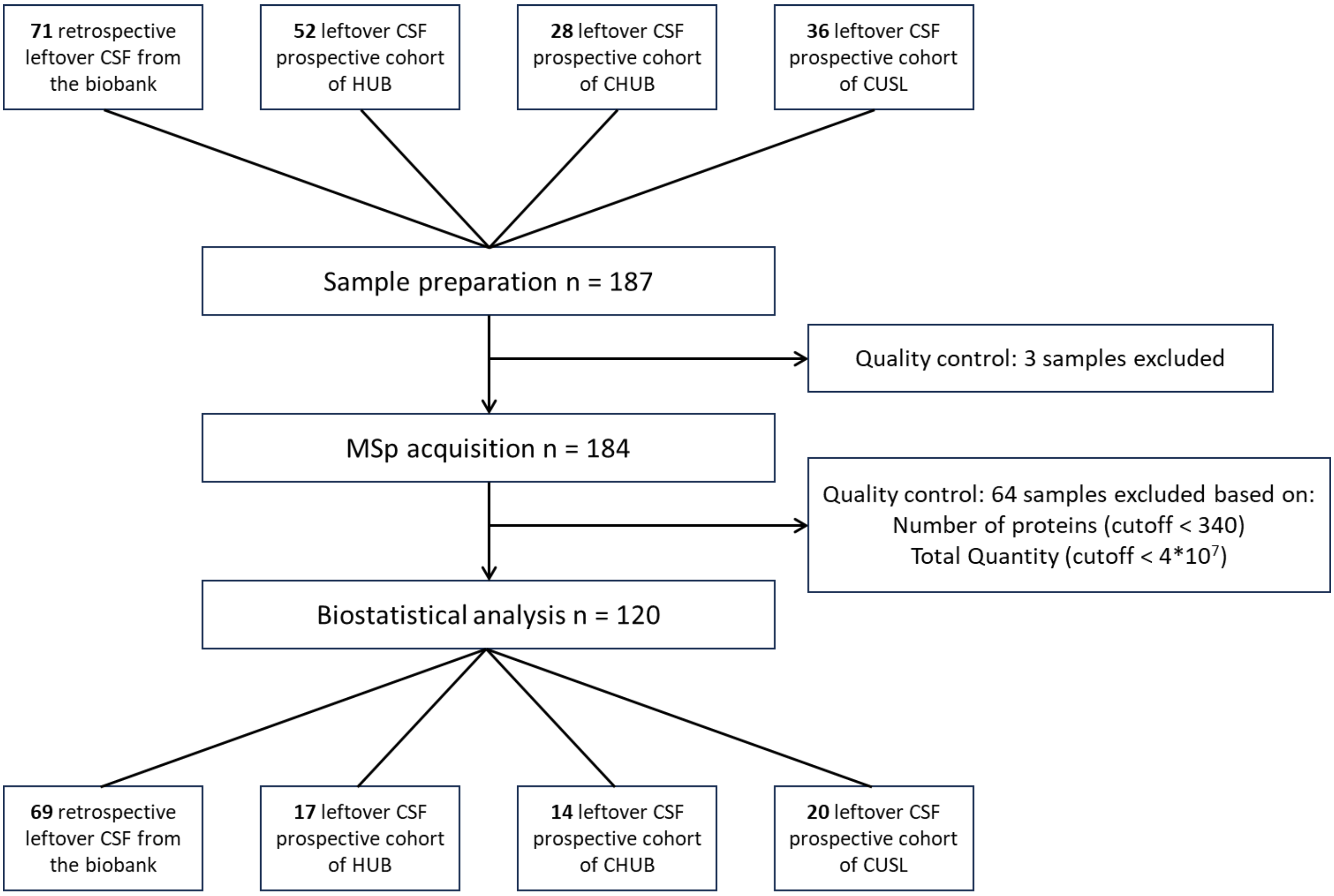


**Supplementary Figure S1 :** Flow chart of the samples selection for the biostatistical analysis.

HUB Erasme Hopital Universitaire de Bruxelles, CHUB Centre Hospitalier Universitaire Brugmann, CUSL Cliniques Universitaires de Saint-Luc.
